# Area-Level Infant Mortality Exposure in Early Life and Later Life Stroke: The Role of Modifiable Risk Factors and Place of Birth

**DOI:** 10.1101/2025.03.27.25324792

**Authors:** Michael Topping, Jason Fletcher

**Affiliations:** Department of Sociology, University of Wisconsin-Madison, Madison, WI, USA; Center for Demography of Health and Aging, University of Wisconsin-Madison, Madison, WI, USA; Center for Demography and Ecology, University of Wisconsin-Madison, Madison, WI, USA; La Follette School of Public Affairs, University of Wisconsin-Madison, Madison, WI, USA; Department of Population Health, University of Wisconsin-Madison, Madison, WI, USA

## Abstract

A growing literature suggests that early life conditions may contribute to the risk of later life stroke. In this paper we explore the role that infant mortality (as a proxy for environmental and socioeconomic conditions) plays in later life self-reported stroke and stroke mortality, while also considering other contextual factors such as state of birth. We use a sample of 244,041 individuals aged 60 and above with the NIH-AARP Diet and Health Study with 16 years of mortality follow-up, allowing us to examine the role that infant mortality rates, along with other known risk factors, play in one’s individual risk of stroke mortality and self-reported stroke. We show using logistic regression models for stroke incidence and cox proportional hazard models for stroke mortality, that infant mortality rates are associated with later life stroke mortality among males, net of controls for education, and stroke risk factors, and with self-reported stroke among females, prior to controlling for risk factors. However, upon controlling for state of birth, these associations dissipate. Our findings suggest that early life factors in the form of infant mortality are important considerations when studying the risk of stroke death for males, for reporting a history of strokes among females and how failure to consider these early life contextual influences may overstate these associations with morbidity and mortality.

## Introduction

Cerebrovascular disease, better known as a stroke, is one of the leading underlying causes of death in the United States, being directly responsible for over 150,000 deaths in 2019 and increasing to over 160,000 in 2020 (Ahmad et al. 2021; Xu et al. 2021). Despite this rise in the crude number of deaths, stroke mortality as a whole has declined over the decades, undoubtedly due to improvements to public health and medicine (Koton et al. 2014). Nevertheless, it is estimated that nearly 800,000 individuals in the United States have a stroke each year, whether new or recurrent, with nearly a fifth of those resulting in death (Benjamin et al. 2017). Thus, reducing both the rate of incidence and the overall mortality of stroke remains a challenge to population health in the United States.

Life course researchers often cite the importance of early-life conditions and how they relate to poor health and mortality at older ages, in addition to more contemporaneous conditions (Lynch and Smith 2005). These early-life conditions come in various forms such as social mobility (Heshmati et al. 2017), intelligence (Backhouse et al. 2015), and place of birth (Topping, Fletcher, and Kim 2024; Topping, Kim, and Fletcher 2021b; Xu et al. 2020; W. Xu, Topping, and Fletcher 2021) to help explain how death and disease operates across the lifespan. One notable thread of work has looked at early-life conditions in the form of infant mortality rates. Infant mortality, or the deaths among children under one year of age, is a powerful metric for a societies overall health (Reidpath 2003). More importantly, it often reflects the social and physical conditions, and especially the disease environment, in which the births occurred.

Indeed, one framework that explains why infant mortality is a great representation for early life socioeconomic and environmental conditions is known as the “Barker hypothesis”. The Barker hypothesis essentially is the idea that higher levels of infant mortality are a result of adverse socioeconomic and nutritional environments in early life (Barker 1991, 1995; Barker and Osmond 1986). In this situation, while many infants die as a result of the adverse conditions, but the survivors who did not likely are scarred by the adversity, which in turn increased both the susceptibility to poor infant outcomes and complications later in life that could result in heart disease and stroke in later life as well among those who survived (Edwards 2019).

Few studies have looked into the role of area-level infant mortality rates as a proxy variable for early-life disease and socioeconomic environments (Leon and Davey Smith 2000). Regarding stroke death in particular, there are only a handful of studies that look exclusively at this relationship between infant mortality and later life stroke. More importantly, when this relationship is examined, it typically is done so by looking across multiple nations or within a European context, with prior research finding that infant mortality rates among birth cohorts in late 1800’s Denmark, England, Finland, France, Netherlands, Norway, and Sweden all saw positive associations with stroke decades later (Amiri et al. 2006a). While informative and robust, this type of relationship has not been thoroughly examined in the context of the United States.

There also is also a lack of consideration to smaller levels of geography, such as the state-level. This is a notable limitation, given that states tend to encompass physical, socioeconomic, and policy environments. As such, individuals born into these places are exposed to these factors both in-utero and after birth, and may have been imprinted differential levels of advantage during a period of critical development. Moreover, not examining states early life conditions in relation to fatal and non-fatal stroke is puzzling given that there is an area of the United States referred to as the “stroke belt” (Lanska and Kuller 1995; Liao et al. 2009). Thus, when opting to look into the relationship between early-life factors and later life stroke, one must factor in place into the analysis, such as state of birth. Indeed, exploring the impact of place of birth onto health outcomes has increasing been used in research (Taylor 2010), due to the fact that one’s state of birth encapsulates many different contextual elements (Xu et al. 2020). In research that looks at stroke or stroke mortality, place of birth in high stroke areas or birth in the stroke belt itself has increasingly found associations with later life disease and mortality (Gilsanz et al. 2017; Glymour et al. 2011; Topping, Kim, and Fletcher 2021a).

Additionally, it is also imperative to consider potential risk factors over time as well. A robust literature has examined modifiable risk factors for stroke across the life course, specifically paying attention to the role of hypertension (Kjeldsen et al. 2017), obesity (Guo et al. 2016), diabetes (Chen, Ovbiagele, and Feng 2016), smoking (Chen et al. 2019), and self-rated health (Mavaddat et al. 2016). Other studies have focused upon educational attainment as another pathway through which stroke incidence and mortality operate, which find that low levels of education are associated with increased risk of mortality and recurrent stroke, along with other cardiovascular events, net of other established risk factors (Che et al. 2020; Cox et al. 2006; Qureshi et al. 2003). Furthermore, the state environments that individuals are born into may shape individuals’ risk and resilience for these risk factors as well.

Finally, it is also critical to consider sex-based differences when it comes to health and mortality in later life, especially as it relates to potential cardiovascular events such as stroke. This is because there has been a noticeable sex gap in stroke incidence and mortality, with males having both higher incidence and mortality from stroke (Giroud et al. 2017; Petrea et al. 2009; Reeves et al. 2008). Yet, more females experience stroke each year, on average, which is often a consequence of their longer life expectancies than their male counterparts. Furthermore, many of the modifiable risk factors for stroke such as smoking and obesity, tend to have a gendered gradient, with males tending to have higher levels of smoking (Higgins et al. 2015) and obesity (Cooper et al. 2021) compared to females, which in turn put them at a higher risk for both incidence and mortality.

Taken together, it is clear that there is a need to consider how area-level infant mortality rates, serving as a proxy for early-life disease and socioeconomic conditions, influence stroke incidence and mortality. Furthermore, there is a need to consider specific risk factors to see if they explain away any potential relationship, as evidenced in prior literature (Avan et al. 2019; Kjeldsen et al. 2017). Contextual factors such as state of birth are also important mechanisms to consider unpacking this relationship with a leading cause of death in the United States. Thus, we address the following questions: 1) is there an association between infant mortality and later life stroke mortality or self-reported stroke? 2) are these associations explained by modifiable risk factors? and 3) how does state of birth influence these effects, if at all? 4) how do these results differ by sex?

## Methods

### Data

This study uses data from the NIH-AARP Diet and Health Study (DHS). The DHS is a study undertaken from 1995 to 1996 in which individuals from the ages of 50-71 years old were recruited from the American Association of Retired Persons (AARP), who responded to a mailed questionnaire (Park et al. 2011; Schatzkin et al. 2001). This questionnaire, initially mailed to 3.5 million members of the AARP, eventually resulted in over 620,000 responses from individuals. From this, nearly 570,000 individuals provided enough information that was compatible for the study of health. The participants of this large study were drawn from six states in the United States (California, Florida, Louisiana, New Jersey, North Carolina, and Pennsylvania), and two metropolitan areas (Atlanta, Georgia and Detroit, Michigan), who provided both written and informed consent. In the initial study in 1995-1996, participants of the DHS were asked information in the questionnaire that revolved around nutrition, along with questions on health, such as prior diagnoses of illness, among others. In addition to the information collected at baseline, basic demographics of each individual (race/ethnicity, sex, etc.) were collected, along with other variables that are commonplace to the study of health and mortality.

### Measures

*Stroke Incidence and Mortality*. The key outcomes in this study are self-reported (non-fatal) stroke, and stroke mortality. Stroke incidence is primarily recorded in the DHS as either a “yes” or “no” response to whether or not the individual has had a history of stroke. Stroke mortality, like other forms of death in the DHS, is measured through death records from annual linkage of the members of the DHS cohort to the Master Death Files of the Social Security Administration (Etemadi et al. 2017). From this, cause of death information was then followed up with searches of the National Death Index (NDI) and corresponding International Classification of Disease (ICD) Codes. The specific codes we use for stroke mortality are I60 through I69. All causes of death in the DHS, including stroke mortality, are provided from follow-up with the DHS through the year 2011, a sixteen year period in total.

*State of Birth*. State of birth is assessed in this study via linkage of individual social security numbers. In the baseline survey, the DHS collected individual social security numbers of respondents. Using the first three digits of these social security numbers, the identification of states of birth is then ascertained (Fletcher 2015).

*Area Level Infant Mortality Rates.* Infant mortality rates are utilized as a proxy for early-life conditions, as done so in prior studies on health and mortality (Amiri et al. 2006b; Barker and Osmond 1986; Leon and Davey Smith 2000; Topping, Kim, and Fletcher 2023). They are calculated at the state level for each year of birth by taking the number of infant deaths over the total number of births of the state, multiplied by 1,000, to acquire the rate. These values are then standardized for the whole sample to have a mean of 0 and a standard deviation of 1. These measures are merged with the NIH-AARP individual level data at the birth year X state level. We obtained these infant mortality rates in a similar manner that other studies have done (Bhalotra and Venkataramani 2011; Topping et al. 2023), which are from volumes of the United States Vital Statistics for the birth years of interest (1925-1937) and for the whole population of the state.

*Covariates.* Age is represented in this study by using each individual’s year of birth. This covariate is included in models to account for the differential risk in both the incidence and mortality of chronic illnesses such as stroke. Sex is also utilized as a way of looking at stroke risk between men and women. Regarding specific risk factors that contribute to stroke, we include measures such as if the individual has a history of heart issues and diabetes, both of these measures are reported as “yes” or “no”. Lifetime smoking status is also included as a control as a risk factor contributing to stroke risk. We also include body mass index (BMI) as a continuous variable to see the potential impact of obesity on stroke. Furthermore, self-rated health is included as a variable (excellent, very good, good, fair, and poor). Finally, educational attainment is included to account for the influence that education has across the lifespan with regard to outcomes such as stroke (Hayward et al. 2021).

### Analytic Strategy

First, descriptive statistics of the DHS sample used in our study are presented. Next, for stroke incidence we opt to employ a series of logistic regression models predicting the individual likelihood of self-reported stroke. We opt to employ a logistic regression model for this outcome due to the fact that we do not have data about when they had the aforementioned stroke, or the frequency of stroke. Regarding stroke mortality, we employ a series of cox proportional hazard models with age as the timescale. For modeling of both self-reported stroke and stroke mortality, Model 1 includes the area-level infant mortality rate (standardized), race, and year of birth as baseline covariates, with model 2 adding in education. Models 3 then controls for specific risk factors (heart disease, diabetes, smoking status, and BMI), in addition to the baseline controls. Finally, Model 4 includes self-rated health at the baseline interview as a covariate. After this, to demonstrate the impact that early-life place plays in health, all models are re-estimated adding state of birth fixed effects. In using state fixed effects, this helps greatly reduce the chance that the relationship between a IMR and stroke is not being driven by potential omitted variables. All analyses are stratified by sex to consider the differences in outcomes that exist between males and females.

## Results

Table 1 presents our sample selection process. From the original DHS sample of 566,398, we remove 165,917 observations that do not have state of birth information or have births in United States territories. Next, we further remove an additional 131,831 observations to exclude those younger than 60 at baseline, and a final 24,609 observations with missing data on covariates, leaving us with a final analytic sample of 244,041. Missing data on covariates was handled using both listwise deletion and multiple imputation, which yielded identical results. As such, results presented below use the former. Sex-stratified summary statistics are presented in Table 2, which report p-values of bivariate associations.. Of the 244,041 observations in our sample, 4,522 males and 1,928 females had a self-reported stroke prior to the baseline survey and 2,222 males and 1,207 females died from a stroke during the 16 year mortality follow up. Over 90% of individuals in the sample were non-Hispanic white, whereas those who were not non-Hispanic white accounted for 7.76% (8,774) of the female sample and 5.68% (6,954) of the male sample. For infant mortality rates (IMR), the average is 62,11 per 1,000 for males and 61,80 for females; however, in our models, we use standardized values of IMR. Regarding risk factors for stroke, 20.86% (32,208), 11.00% (16,985), and 71.09% (109,766) of male respondents reported a self-reported history of heart disease, diabetes, and having ever smoked, respectively. Whereas female’s respondents reported 10.31% (9,240), 8.16% (7,318), and 54.81% (49,134). For BMI, the average among respondents was 27.03 for males and 26.69 for females. Given our large sample size there may be concern about our minimum significance threshold we adopt (p<0.05); however, we remove a substantial amount of variation through year of birth and state of birth fixed effects, so our effective sample size is much smaller. As such, we believe using this standard threshold is appropriate in this situation.

**Table 1.** Study Sample Selection.

| Sample Size | Removed | Reason? |
| --- | --- | --- |
| Original Sample (N=566,398) | 165,917 | Missing state of birth information or born in United States territories |
| 400,481 | 131,831 | Younger than 60 years of age at baseline |
| 268,650 | 24,609 | Missingness on other variables |
| Final Sample (N=244,041) |  |  |

**Table 2.**
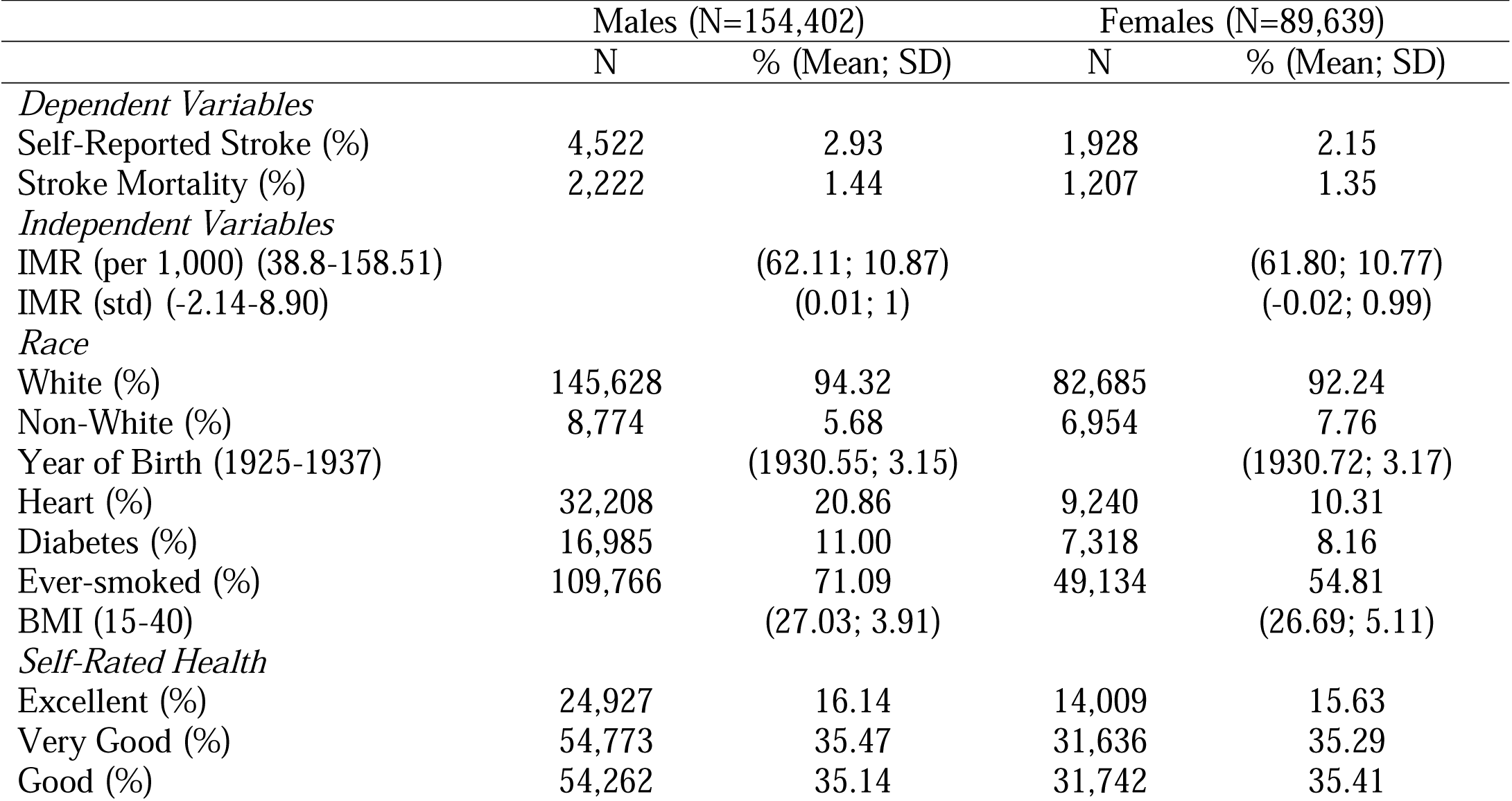
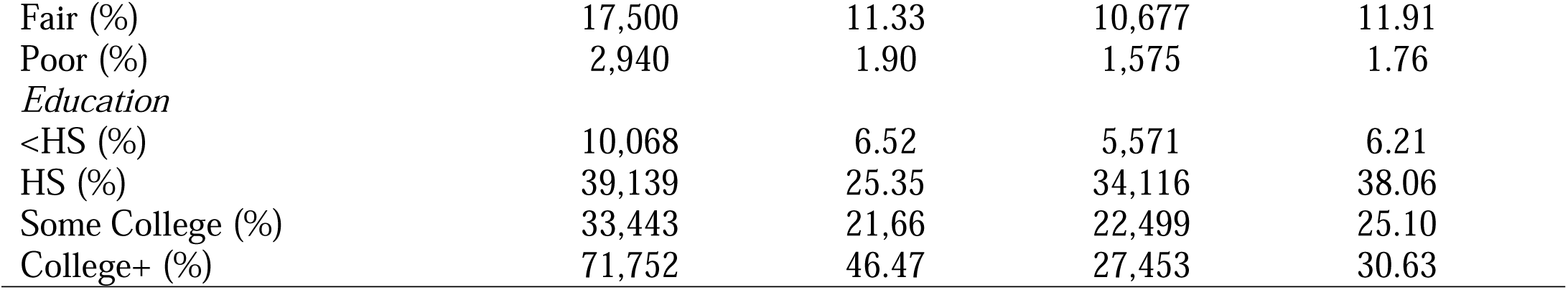
Descriptive Statistics by Sex.

|  | Males (N=154,402) |  | Females (N=89,639) |  |
| --- | --- | --- | --- | --- |
|  | N | % (Mean; SD) | N | % (Mean; SD) |
| *Dependent Variables* |  |  |  |  |
| Self-Reported Stroke (%) | 4,522 | 2.93 | 1,928 | 2.15 |
| Stroke Mortality (%) | 2,222 | 1.44 | 1,207 | 1.35 |
| *Independent Variables* |  |  |  |  |
| IMR (per 1,000) (38.8-158.51) |  | (62.11; 10.87) |  | (61.80; 10.77) |
| IMR (std) (-2.14-8.90) |  | (0.01; 1) |  | (-0.02; 0.99) |
| *Race* |  |  |  |  |
| White (%) | 145,628 | 94.32 | 82,685 | 92.24 |
| Non-White (%) | 8,774 | 5.68 | 6,954 | 7.76 |
| Year of Birth (1925-1937) |  | (1930.55; 3.15) |  | (1930.72; 3.17) |
| Heart (%) | 32,208 | 20.86 | 9,240 | 10.31 |
| Diabetes (%) | 16,985 | 11.00 | 7,318 | 8.16 |
| Ever-smoked (%) | 109,766 | 71.09 | 49,134 | 54.81 |
| BMI (15-40) |  | (27.03; 3.91) |  | (26.69; 5.11) |
| *Self-Rated Health* |  |  |  |  |
| Excellent (%) | 24,927 | 16.14 | 14,009 | 15.63 |
| Very Good (%) | 54,773 | 35.47 | 31,636 | 35.29 |
| Good (%) | 54,262 | 35.14 | 31,742 | 35.41 |
| Fair (%) | 17,500 | 11.33 | 10,677 | 11.91 |
| Poor (%) | 2,940 | 1.90 | 1,575 | 1.76 |
| *Education* |  |  |  |  |
| <HS (%) | 10,068 | 6.52 | 5,571 | 6.21 |
| HS (%) | 39,139 | 25.35 | 34,116 | 38.06 |
| Some College (%) | 33,443 | 21.66 | 22,499 | 25.10 |
| College+ (%) | 71,752 | 46.47 | 27,453 | 30.63 |

Table 3 presents odds ratios for self-reported strokes, also stratified by sex. For males, it is shown that IMR is not associated with the odds of self-reported stroke across all models once factors such as education are accounted for. On the other hand, IMR is associated with a 7% odds increase in self-reported stroke amongst females (OR=1.07; CI: 1.01-1.13; p < 0.043) when controlling for race and year of birth. Being non-white is associated with higher odds of self-reported stroke (OR=1.38; CI: 1.19-1.60; p < 0.001). In Model 2 for females, IMR predicts a 6% increase in the likelihood of self-reported stroke (OR=1.06; CI: 1.00-1.11; p < 0.047), with this association dissipating when factoring in modifiable risk factors and self-rated health. This suggests that potentially IMR in early life influences the risk of stroke indirectly, operating through known-risk factors for stroke. We also opted to re-run the analysis controlling for state of birth fixed effects. Upon adding in state of birth fixed effects, the associations for IMR lost statistical significance (see Online Supplement for more details).

**Table 3.** Odds Ratios Predicting the Likelihood of Self-Reported Stroke.

|  | Males |  |  |  | Females |  |  |  |
| --- | --- | --- | --- | --- | --- | --- | --- | --- |
|  | Model 1 | Model 2 | Model 3 | Model 4 | Model 1 | Model 2 | Model 3 | Model 4 |
| IMR (std) | 1.04* | 1.03 | 1.03 | 1.00 | 1.07* | 1.06* | 1.03 | 0.99 |
|  | (1.01 - 1.08) | (0.99 - 1.07) | (0.99 - 1.07) | (0.96 - 1.04) | (1.01 - 1.13) | (1.00 - 1.11) | (0.98 - 1.09) | (0.94 - 1.05) |
| Non-White | 1.09 | 1.07 | 1.10 | 0.99 | 1.38*** | 1.35*** | 1.25** | 1.09 |
|  | (0.97 - 1.24) | (0.94 - 1.21) | (0.97 - 1.25) | (0.87 - 1.12) | (1.19 - 1.61) | (1.16 - 1.57) | (1.07 - 1.46) | (0.94 - 1.28) |
| Birth Year | 0.93*** | 0.93*** | 0.95*** | 0.94*** | 0.95*** | 0.96*** | 0.96*** | 0.96*** |
|  | (0.92 - 0.94) | (0.92 - 0.95) | (0.94 - 0.96) | (0.93 - 0.96) | (0.94 - 0.97) | (0.94 - 0.97) | (0.95 - 0.98) | (0.95 - 0.98) |
| *Risk Factors* |  |  |  |  |  |  |  |  |
| Heart Disease |  |  | 2.56*** | 1.52*** |  |  | 3.81*** | 2.23*** |
|  |  |  | (2.41 - 2.73) | (1.42 - 1.62) |  |  | (3.44 - 4.22) | (2.01 - 2.48) |
| Diabetes |  |  | 2.08*** | 1.40*** |  |  | 2.05*** | 1.38*** |
|  |  |  | (1.93 - 2.24) | (1.30 - 1.51) |  |  | (1.82 - 2.32) | (1.22 - 1.56) |
| Eversmoked |  |  | 1.40*** | 1.21*** |  |  | 1.31*** | 1.18*** |
|  |  |  | (1.30 - 1.51) | (1.12 - 1.30) |  |  | (1.20 - 1.44) | (1.08 - 1.30) |
| BMI |  |  | 0.99* | 0.97*** |  |  | 1.02*** | 0.99 |
|  |  |  | (0.98 - 1.00) | (0.97 - 0.98) |  |  | (1.01 - 1.03) | (0.98 - 1.00) |
| *Self-Rated Health* |  |  |  |  |  |  |  |  |
| (ref. Excellent) |  |  |  |  |  |  |  |  |
| Very Good |  |  |  | 1.82*** |  |  |  | 2.00*** |
|  |  |  |  | (1.53 - 2.16) |  |  |  | (1.49 - 2.69) |
| Good |  |  |  | 4.23*** |  |  |  | 5.38*** |
|  |  |  |  | (3.60 - 4.98) |  |  |  | (4.07 - 7.12) |
| Fair |  |  |  | 10.76*** |  |  |  | 11.91*** |
|  |  |  |  | (9.1 - 12.7) |  |  |  | (8.95 - 15.9) |
| Poor |  |  |  | 19.11*** |  |  |  | 21.56*** |
|  |  |  |  | (15.8 - 23.1) |  |  |  | (15.7 - 29.6) |
| *Education* (ref. <HS) |  |  |  |  |  |  |  |  |
| High School |  | 0.81*** |  |  |  | 0.58*** |  |  |
|  |  | (0.73 - 0.91) |  |  |  | (0.50 - 0.68) |  |  |
| Some College |  | 0.76*** |  |  |  | 0.58*** |  |  |
|  |  | (0.68 - 0.85) |  |  |  | (0.49 - 0.68) |  |  |
| College+ |  | 0.52*** |  |  |  | 0.40*** |  |  |
|  |  | (0.47 - 0.58) |  |  |  | (0.34 - 0.47) |  |  |
| Observations | 154,402 | 154,402 | 154,402 | 154,402 | 89,639 | 89,639 | 89,639 | 89,639 |

Table 4 presents hazard ratios for stroke mortality, stratified by sex. For males in Model 1, IMR is shown to be an important predictor in the risk of stroke mortality, with each standard deviation increase in IMR being associated with a 9% greater risk of death (HR=1.09; CI: 1.04-1.14; p < 0.001), net of controls for race and year of birth. Model 2 includes controls for education, which maintains the association between IMR and stroke mortality, and presents an 8% greater risk of death (HR=1.08; CI: 1.03-1.13; p < 0.001). Upon the introduction of risk factors in Model 3, all, with the exception of BMI, is associated with both males and females’ risk of stroke mortality. However, the relationship between IMR and stroke mortality remains for males (HR=1.08; CI: 1.0-1.14; p < 0.001), but not females (HR=0.97; CI: 0.90-1.04; p < 0.385). Finally, Model 4 controls for self-rated health in addition to previous risk factors, and is shown that as both male and females report worse health, their risk of stroke mortality increases. Moreover, as with prior models, the association between IMR and stroke mortality remains for males (HR=1.07; CI: 1.02-1.12; p < 0.005), but not females (HR=0.96; CI: 0.89-1.03; p < 0.135), with males’ risk of stroke increasing 7% on average for each standard deviation increase in IMR.

**Table 4.**
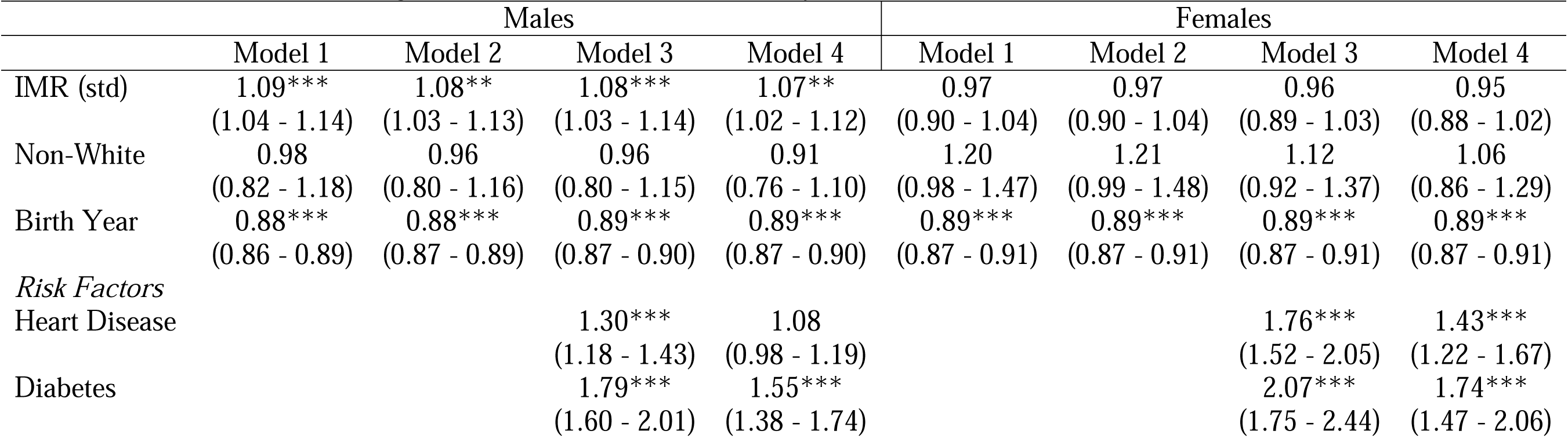
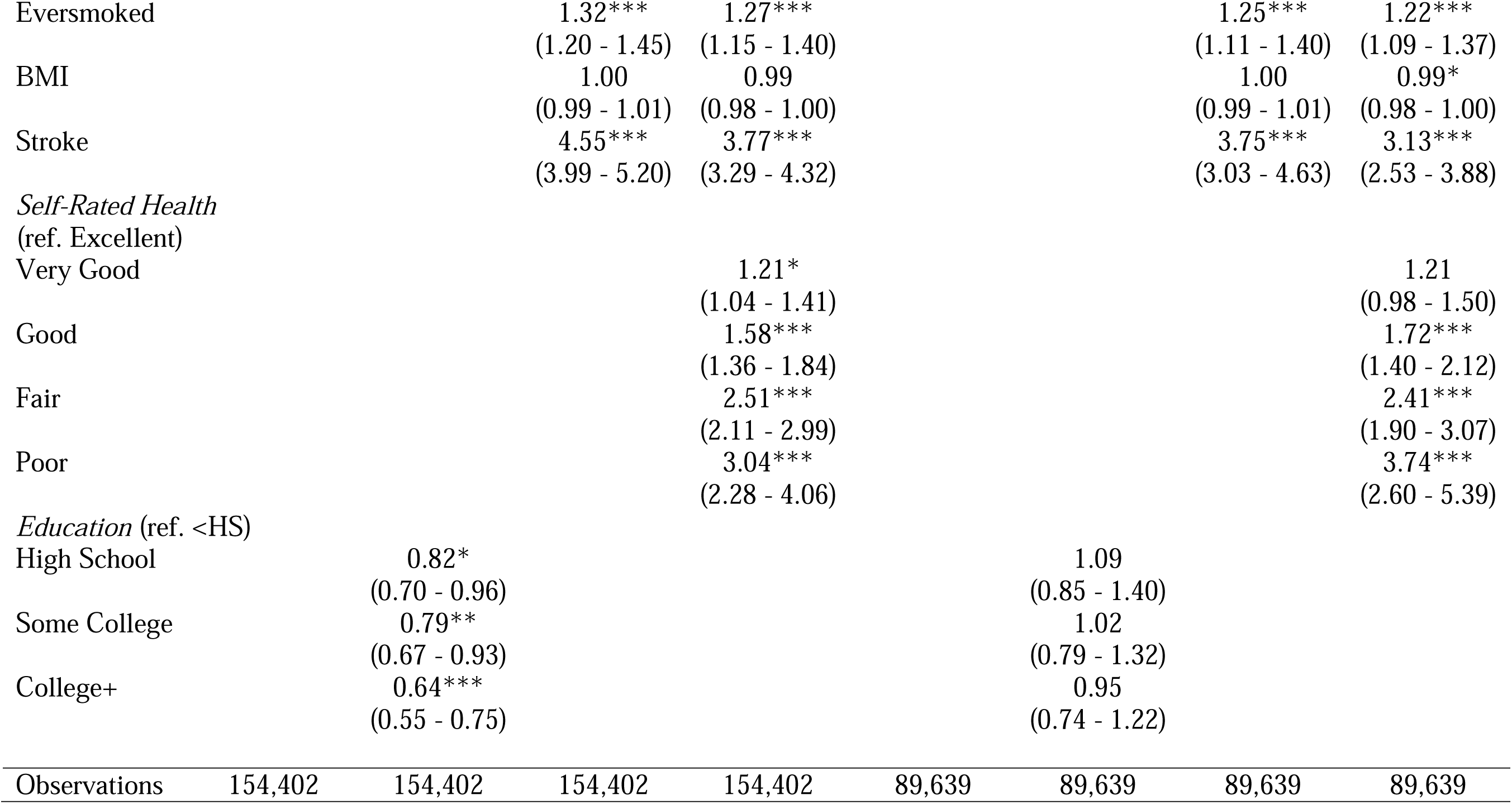
Hazard Ratios Predicting the Likelihood of Stroke Mortality.

## Discussion

Most research concerning stroke incidence and stroke mortality has emphasized individual level factors. Meanwhile, research that examines contextual factors on forms of health and mortality has grown in recent years, especially for cardiovascular disease (Alastalo et al. 2013; Murray et al. 2014), but not stroke. Our study contributes to the examination of early-life factors by exploring the impact that infant mortality has on later life fatal and non-fatal stroke. Whereas prior studies often are concerned with factors such as birthweight (Class et al. 2014), we opt to use infant mortality as a useful proxy for early-life contextual conditions, along with controls for geographic contexts via state of birth. Ultimately, the findings presented in this paper suggest several takeaways.

First, we further the understanding of the examination of sex based differences in stroke survival and mortality. Our results indicate that higher IMR leads to an increased risk of stroke mortality among men, even when controlling for elements such as known risk factors for stroke such as diabetes and smoking status. For females, our results lend some support to the notion that higher IMR leads to an increased likelihood for reporting a history of stroke. This is consistent with prior examinations of morbidity and mortality, where researchers have found that men have a higher likelihood of death from chronic disease, whereas women are likely to live with them longer as a result of their longer life expectancies (Case and Paxson 2005). Furthermore, these results are consistent with the Barker hypothesis, or the idea that exposure to adverse nutritional and socioeconomic conditions during a critical period of development can imprint a disadvantage in early life that accumulates into poor health later on. Future work should further delve into this phenomenon regarding other causes of death, to see if it is consistent with specific types of death, or if it is proven true more broadly.

Second, we find that when state of birth fixed effects are included in our estimations of self-reported stroke and stroke mortality, that their association with IMR dissipates, but other mechanisms that are factored in remain relatively the same. This implies IMR measures capture broader state-level factors such as different policy regimes (i.e. minimum wage, earned income tax credit) or resource access in the form of healthcare, which have important implications for health and mortality across the life course. Failure to control for these factors inflate associations between early-life IMR and stroke outcomes. The state environments people were born into may therefore have an impact through various mechanisms that work to influence modifiable risk factors later in the life course, which in turn are associated with self-reported stroke and stroke mortality.

This research, like any, has limitations that are in need of being addressed in future work. First, the most notable limitation to this study is the fact that one of our outcomes, self-reported stroke, is both a self-reported measure and lacks information on when the individual had a stroke, or how many, and lacks adjudication. Future research may examine measures that better gauge the timing and the quantity of stroke episodes that an individual has throughout their lifetime. Related to this, there is the notion of misattribution with regards to stroke mortality. This is because at times, there are problems of attributing stroke as a cause of mortality, due to how it is classified as an outcome of death (Lackland et al. 2014; Viitanen, Winblad, and Asplund 2009). However, we believe this to not be a significant issue that alters the findings of this paper. Second, the data utilized in this study was gathered from individuals living across eight states within the United States, thus limiting its geographic coverage. Given that chronic illnesses such as stroke have higher incidence and mortality in specific states and regions of the United States, future research should look to include information from individuals across all fifty states. We do note that while the *current* residence of respondents was constrained to these eight places, respondents were born in all fifty states, so we are able to capture national variation in IMR during this period. Yet, given that the individuals born during this time period (1920s to 1940s) may or may not have acquired social security numbers as young adults rather than as infants, the first 3 digits of the social security may not capture this as robustly. Future studies should look into examining the associations of this paper with data with complete geographic coverage of the United States.

Another limitation to be noted is the methods in examining IMR and stroke in a broader context. Better approaches to the study of infant mortality would better help situate both the degree and severity of specific socioeconomic and environmental conditions, such as the age of infant death (i.e. one day vs eleven months), the cause of death, and the seasonal time of death, to name a few. Adding this layer to the examination of the relationship between IMR and later life health in future studies would better articulate the impact that adverse conditions across the life course. Finally, the representativeness of the sample is also a limitation to consider. Given that 63.1% of our sample is male and 36.9% is female, the results should not be taken as representative of the whole United States. However, it should be noted that despite these issues of representativeness, our paper does reaffirm prior findings on early life factors and old age health and mortality.

## Conclusion

Despite the above limitations, this research is unique in the sense that our data allows us to explore instances of stroke that are both fatal and non-fatal. This enables us to examine the extent by which early-life factors, along with mechanisms such as education and self-rated health, contribute to differences in risk of late life stroke by sex. Ultimately, our results suggest that early-life disease and socioeconomic conditions, as measured by exposure to area-level infant mortality rates, on average, are associated with increased risk of stroke death for males, and females for reporting a history of stroke. However, upon accounting for state of birth in the analysis, these associations then dissipate, implying the need to consider early life contextual factors in the examination of health and mortality. Future studies would benefit from employing larger sample sizes, particularly for non-white populations, and incorporating additional measures that capture structural early-life conditions, such as air pollution, which have been shown to program poor fetal development (Burris and Baccarelli 2017). Ascertaining which particular early-life factors matter more regarding the risk for both stroke incidence and mortality would help to better inform various public health interventions in the United States, and in turn improve overall population health.

## Data Availability

All data produced in the present work are restricted, given the use of geographic identifiers.

## Notes

### Competing Interest Statement

The authors have declared no competing interest.

### Funding Statement

This research was carried out using the facilities of the Center for Demography and Ecology and Center, supported by Eunice Kennedy Shriver National Institute of Child Health and Human Development grant P2C HD047873 and the Center for Demography of Health and Aging, which is supported by National Institute on Aging grant P30 AG017266. Topping also received support by the National Institute on Aging training grant T32 AG000129.

### Author Declarations

This study received approval from the University of Wisconsin-Madison Institutional Review Board.

